# Urban greenspace and anxiety symptoms during the COVID-19 pandemic: A 20-month follow up of 19,848 participants in England

**DOI:** 10.1101/2022.04.27.22274371

**Authors:** Feifei Bu, Hei Wan Mak, Andrew Steptoe, Benedict W Wheeler, Daisy Fancourt

**Affiliations:** Department of Behavioural Science and Health, Institute of Epidemiology & Health Care, University College London; European Centre for Environment and Human Health, University of Exeter Medical School

**Keywords:** greenspace, anxiety, COVID-19, nature, panel data, growth curve modelling

## Abstract

This study examined the association between greenspace and the growth trajectories of anxiety symptoms during the COVID-19 pandemic. Using data from 19,848 urban residents in England who were followed for 20 months between March 2020 and October 2021, we found that living in an area with higher greenspace coverage was associated with fewer anxiety symptoms over time independent of population density, area deprivation levels, socio-demographics, and health profiles. There was limited evidence that greenspace was related to the change of anxiety symptoms over time. No association with anxiety trajectories was found when using greenspace proximity.

## Introduction

There is a growing body of literature on the effect of exposure to natural environment or greenspace on mental health.^1–4^ For example, a Dutch study that used primary care records linked to greenspace in people’s postcode areas found that a higher percentage of greenspace was associated with lower odds of depressive and anxiety disorders.^5^ Another large-scale observational study in Australia found that a higher exposure to greenspace was associated with lower incidence of psychological distress.^6^ Further, a German survey that followed the same participants over 12 years found that living a shorter distance to urban greenspace was associated with higher life satisfaction.^7^ Similar findings have also been found in experimental studies. A study in Japan reported that exposure to a forest environment significantly decreased feelings of hostility and depression compared to a control day (not visiting a forested area).^8^ And a randomised trial in the United States (US) found that feelings of depression and worthlessness were significantly lower in the greening intervention group (cleaning and greening vacant lots) compared to the control group without any intervention.^9^

Several theories have been proposed to understand the mechanisms of the psychological benefits of greenspace. The widely recognised theories include (but are not limited to) the attention restoration theory (ART), stress reduction theory (SRT), and neighbourhood effect theory. ART suggests that natural environment exposure enables recovery from directed attention fatigue due to prolonged engagement in mental-demanding tasks.^10^ Similarly, SRT suggests that exposure to nature can activate a quick positive affective response and initiate the restorative process because it provides a breather from stress or blocks negative thoughts and feelings.^11^ Benefits of nature and greenspace may also arise from the lifestyle and behaviours which people engage in when exposed to greenspace. According to the theory of neighbourhood effect, greenspace may help sustain and improve health by providing venues for physical activities (e.g. walking, running, cycling and gardening), and by facilitating social interaction with a community.^12^ People living in areas with access to greenspace may be more motivated to engage in these activities and interactions, which are known to have positive effects on mental health and wellbeing. These theories situate within a generalised framework of three nature-health pathway domains: restoring capacity (e.g. ART and SRT); building capacity (e.g. physical activity and social contact); and reducing harm (e.g. reducing exposure to noise).^13^

Since December 2019, the world has been devastated by the outbreak of coronavirus disease (COVID-19). Many countries, including the United Kingdom (UK), implemented lockdown or stay-at-home orders to control the spread of the virus. The restrictions that these imposed on individuals’ movements is shown by reductions of 70-90% in the use of public transport and 40-80% in driving and walking during the first national lockdown (23^rd^ March-10^th^ May 2020).^14^ Notably, movements remained below usual levels even after the restrictions were lifted in the UK.^14^ Yet despite this general decrease in movements, there was in fact an increase in park/forest visitations globally during the pandemic compared to before before.^15,16^ This suggests that individuals prioritised opportunities to engage in nature and spend time in greenspace, raising the question as to whether such behaviours were undertaken as part of coping strategies to support and sustain mental health and wellbeing.

Studies have consistently shown adverse effects on mental health during the COVID-19 pandemic across all age groups, in particular in countries where greater social restrictions were imposed.^17,18^ But as yet, the literature on whether greenspace supported mental health in the pandemic is in its early stages, and results are mixed.^19^ Studies that used self-reported measures of nature experience found that nature views from home and accessible greenspace in the neighbourhood were associated with a lower level of depression and anxiety during the pandemic.^20,21^ However, in another study although people who self-reported a decrease in visiting greenspace during the pandemic were found to have a higher risk of major depression, no evidence was found for anxiety.^22^ Similar findings were reported in another study that used tree-rich greenspace measures from residential postcodes.^23^ However, most studies on greenspace and mental health during COVID-19 have used cross-sectional data, focusing on particular short periods during the pandemic (e.g. first lockdown). There is a lack of longitudinal studies looking at how greenspace is related to mental health changes over time.

In this light, this study examined the relationship of greenspace and anxiety during the COVID-19 pandemic. We used data from 25,390 adults living in urban areas in England who were followed for 20 months between March 2020 and October 2021. Data were analysed using latent growth modelling, which allowed us to examine how access to urban greenspace was related to the levels of and rate of change in anxiety across different stages of the COVID-19 pandemic. Our study provided an advance on previous research on greenspace and mental health in several ways. First and methodologically, a challenge in pre-pandemic observational studies is that residential greenspace data are often assumed to proxy for direct exposure through visits, while it is clear that many people often visit nature some distance from their home.^24^ However, the COVID-19 restrictions, especially strict lockdowns, confined people to their homes and local neighbourhood,^25,26^ providing a unique opportunity to explore the psychological benefits of residential greenspace. Secondly, as mentioned above, this study followed individuals over a longer period through the course of the pandemic compared to most other studies.^19^

More robust evidence, such as this, is important for formulating guidance for people to stay mentally well during and after pandemics. This is particularly timely as it has been estimated that 49.6% of the British population experienced anxiety at the start of the pandemic, with 10 million projected to need mental health support as a direct consequence of the crisis.^27,28^ Moreover, this study has implications for the new UK government pilot study of ‘Green Social Prescribing’ and other nature-based social prescribing schemes to support people’s mental health and wellbeing through greenspace and nature-based activities (e.g. health walks, green gyms, healthy campfires, and community food growing). Finally, the present study will also be valuable to inform long-term policies linking health and natural environment, such as the 25 Year Environment Plan set out by the UK government to help restore and improve the nature, promote health and wellbeing through the natural environment, as well as increasing the public’s awareness of the value of spending time in greenspace.^29^

## Method

### Research design and participants

This study analysed data from the University College London (UCL) COVID-19 Social Study, a large panel study of the psychological and social experiences of over 75,000 adults (aged 18+) in the UK during the COVID-19 pandemic. The study commenced on 21 March 2020 and involved weekly and then monthly (four-weekly) online data collection from participants for the duration of the pandemic. The study did not use a random sample design and therefore the original sample is not representative of the UK population. However, it does contain a heterogeneous sample that was recruited using three primary approaches. First, convenience sampling was used, including promoting the study through existing networks and mailing lists (including large databases of adults who had previously consented to be involved in health research across the UK), print and digital media coverage, and social media. Second, more targeted recruitment was undertaken focusing on (i) individuals from a low-income background, (ii) individuals with no or few educational qualifications, and (iii) individuals who were unemployed. Third, the study was promoted via partnerships with third sector organisations to vulnerable groups, including adults with pre-existing mental health conditions, older adults, carers, and people experiencing domestic violence or abuse. The study was approved by the UCL Research Ethics Committee [12467/005] and all participants gave informed consent. A full protocol for the study is available online at https://osf.io/jm8ra/.

In the present study, we restricted the sample to participants living in England (N= 58,726). Postcodes that were crucial for linking with greenspace access were collected at later stages of the COVID-19 Social Study (February and November 2021). These were available for 30,529 participants who had not moved addresses since March 2020. Further, we excluded participants with fewer than three repeated measures (9.1%) or with missing data on any of the outcome variables or predictors (9.2%). This left us an analytical sample of 24,934 participants who were followed up for a maximum of 20 months from March 2020 to October 2021. Considering the substantive differences in greenspace accessibility and type in rural areas,^30^ this study focused on participants living in urban areas based on the 2011 rural-urban classification for small area geographies (N=12,658). Please see the supplementary Figure S1 for the distribution of the included areas.

### Measurements

#### Anxiety symptoms

Anxiety symptoms were measured using the Generalized Anxiety Disorder assessment (GAD-7)^31^; a well-validated tool used to screen for generalized anxiety disorder in clinical practice and research. These questions were also worded as ‘over the last week’ for the same reason as the depression items. The GAD-7 comprises 7 items with 4-point responses ranging from ‘not at all’ to ‘nearly every day’, with higher overall scores indicating more symptoms of anxiety, ranging from 0 to 21.

#### Greenspace

‘Greenspace’ is used here as a generic term for natural environments broadly, and availability was measured by percentages of natural environment land cover (e.g. woodland, grassland, mountain, heath & bog, arable/horticultural site etc.) within Lower-layer Super Output Area (LSOA). Land cover data were derived from the 2019 UKCEH Land Cover Map.^32^ The 20m raster land cover dataset was intersected with LSOA boundaries using a geographic information system (ArcGIS 10.6, ESRI, Redlands CA), and the percentage area of all land cover types excluding urban/suburban was calculated for each area. LSOAs are small areas designed to be of a similar population size, with an average of approximately 1,500 residents. Participants’ postcodes were linked to their LSOA of residence using the Office for National Statistics Postcode Directory. 12,685 urban LSOAs (out of 26,989 in England) included at least one participant from the analytical sample. The greenspace percentage in the residential area was coded as a categorical variable with four categories: ≤10%, <10-20%, <20-50%, >50%.^33^

In addition to percentages, we used proximity to the closest public parks and playing fields as an alternative greenspace measure. This was obtained from the Office for National Statistics.^34^ We also considered a subjective evaluation of greenspace as part of sensitivity analyses. This was measured by how satisfied people were with the availability of usable greenspace in their neighbourhood (satisfied vs neutral/dissatisfied). This information was collected only once in July 2020, and was available for a reduced number of participants (N=12,570).

#### Covariates

We controlled for two area level factors, population density and area deprivation. Population density was of particular importance in context of COVID-19 which might be directly or indirectly related to anxiety symptoms. Population density was defined as number of people square kilometre, which was obtained from the Office for National Statistics based on LSOA. This was categorised into three categories: ≤2,500, <2,500-5,000, >5,000.

Area deprivation was measured by Index of Multiple Deprivation (IMD, 2019) at LSOA level. This was coded in deciles with 1 being the most deprived and 10 the least deprived. IMD considered seven domains of deprivation, including income, employment, education, health, crime, barriers to housing and services and living environment. The environment domain only included housing condition, outdoor air quality and road traffic accidents, which therefore does not overlap with the greenspace measures.

In addition, we considered a number of individual characteristics as potential confounders. These included gender (women vs men), ethnicity (white vs ethnic minorities), age groups (age 18-29, 30-45, 46-59, 60+), education (up to GCSE levels, A-levels or equivalent, university degree or above), annual income (<£16,000, ≤£16,000-29,999, ≤£30,000-59,999, ≤£60,000-89,999, ≥£90,000), employment status (employed vs other), self-reported diagnosis of any long-term physical health condition (e.g., asthma or diabetes) or any disability (yes vs no), and self-reported diagnosis of any long-term mental health condition (e.g., depression, anxiety) (yes vs no).

### Statistical analysis

Data were analysed using the latent growth modelling (LGM) approach. More specifically, we used piecewise LGM which deals with nonlinear growth trajectories by breaking into separate linear pieces. The choice of knots (break points) was informed by previous research^35,36^ and the data. We started by an unconditionally latent growth model of anxiety symptoms without any predictors, followed by the model with only greenspace to predict the growth factors (intercept and slope) (Model I). Then, we added the area factors, populational density and IMD, to the model (Model II). Finally, the full model additionally controlled for individual characteristics (Model III).

In addition to the main analyses, a sensitivity analysis on a subsample (N=12,570) was conducted to consider both quantity and quality of greenspace exposure. We also tested an alternative approach (free time scores) which makes no assumption about the shape of growth trajectory and allows it to be determined by data. Further sensitivity analyses were carried out using alternative greenspace measures, as well as controlling time-varying covariates (days of going outside the house last week). Weights were applied throughout the analyses. The analytical sample (include rural sample) was weighted to the proportions of gender, age, ethnicity and education in the English population obtained from the Office for National Statistics.^37^ Main analyses were implemented in Mplus Version 8.

## Results

### Descriptive statistics

As shown in Table 1, in the unweighted sample of 19,848 participants, women (75.6%) and people with a university degree or above (69.8%) were overrepresented, whereas younger adults (aged 18-29; 4.5%) and people from ethnic minority groups (4.8%) were underrepresented. After weighting, the sample reflected population proportions, with 52.9% women, 36.8% participants with a degree or above, 17% aged under 30, and 13.8% participants belonging to an ethnic minority group. Approximately 48% participants lived in areas of 10 percent greenspace or less, 14.6% between 10 and 20 percent, another 20.7% between 20 and 50 percent, and 16.6% lived in areas with more than 50 percent of greenspace.

**Table 1.** Sample characteristics (N=19,848)

|  |  | Unweighted | Weighted |
| --- | --- | --- | --- |
| Greenspace | ≤10% | 46.8% | 48.0% |
|  | <10-20% | 14.1% | 14.6% |
|  | <20-50% | 20.9% | 20.7% |
|  | >50% | 18.2% | 16.6% |
| Greenspace | Satisfied | 84.6% | 81.8% |
| Satisfaction (N=12,570) | Neutral/dissatisfied | 15.4% | 18.2% |
| Population density | ≤2,500 | 26.3% | 24.2% |
|  | <2,500-5,000 | 31.6% | 31.2% |
|  | >5,000 | 42.2% | 44.6% |
| IMD decile |  | 6.0 (2.8) | 5.5 (2.8) |
| Gender | Men | 24.4% | 47.1% |
|  | Women | 75.6% | 52.9% |
| Ethnicity | White | 95.2% | 86.2% |
|  | Ethnic minority | 4.8% | 13.8% |
| Age group | 18-29 | 4.5% | 17.0% |
|  | 30-45 | 25.3% | 28.5% |
|  | 46-59 | 34.4% | 24.6% |
|  | 60+ | 35.8% | 29.9% |
| Education | GCSE or below | 13.6% | 32.3% |
|  | A levels or equivalent | 16.6% | 30.9% |
|  | Degree or above | 69.8% | 36.8% |
| Annual income | <£16,000 | 14.2% | 18.6% |
|  | ≤£16,000-29,999 | 24.0% | 26.8% |
|  | ≤£30,000-59,999 | 35.2% | 33.5% |
|  | ≤£60,000-89,999 | 15.7% | 13.4% |
|  | ≥£90,000 | 10.9% | 7.8% |
| Employment status | Not employed | 36.7% | 38.6% |
|  | Employed | 63.3% | 61.4% |
| Physical health condition | No | 58.5% | 58.7% |
|  | Yes | 41.5% | 41.3% |
| Mental health condition | No | 82.9% | 80.2% |
|  | Yes | 17.1% | 19.8% |

### Latent growth models

Figure 1 shows the estimated growth trajectory of anxiety symptoms from the unconditional LGM, together with the stringency index of the strictness of COVID-19 responses in England^38^ and the number of new confirmed COVID-19 cases. Generally, anxiety symptoms decreased over the first national lockdown period and following the easing of restrictions. However, it started to increase around August 2020 and peaked in November when England entered the second national lockdown, before a gradual and slow decrease until the end of the follow-up period.

**Figure 1.**
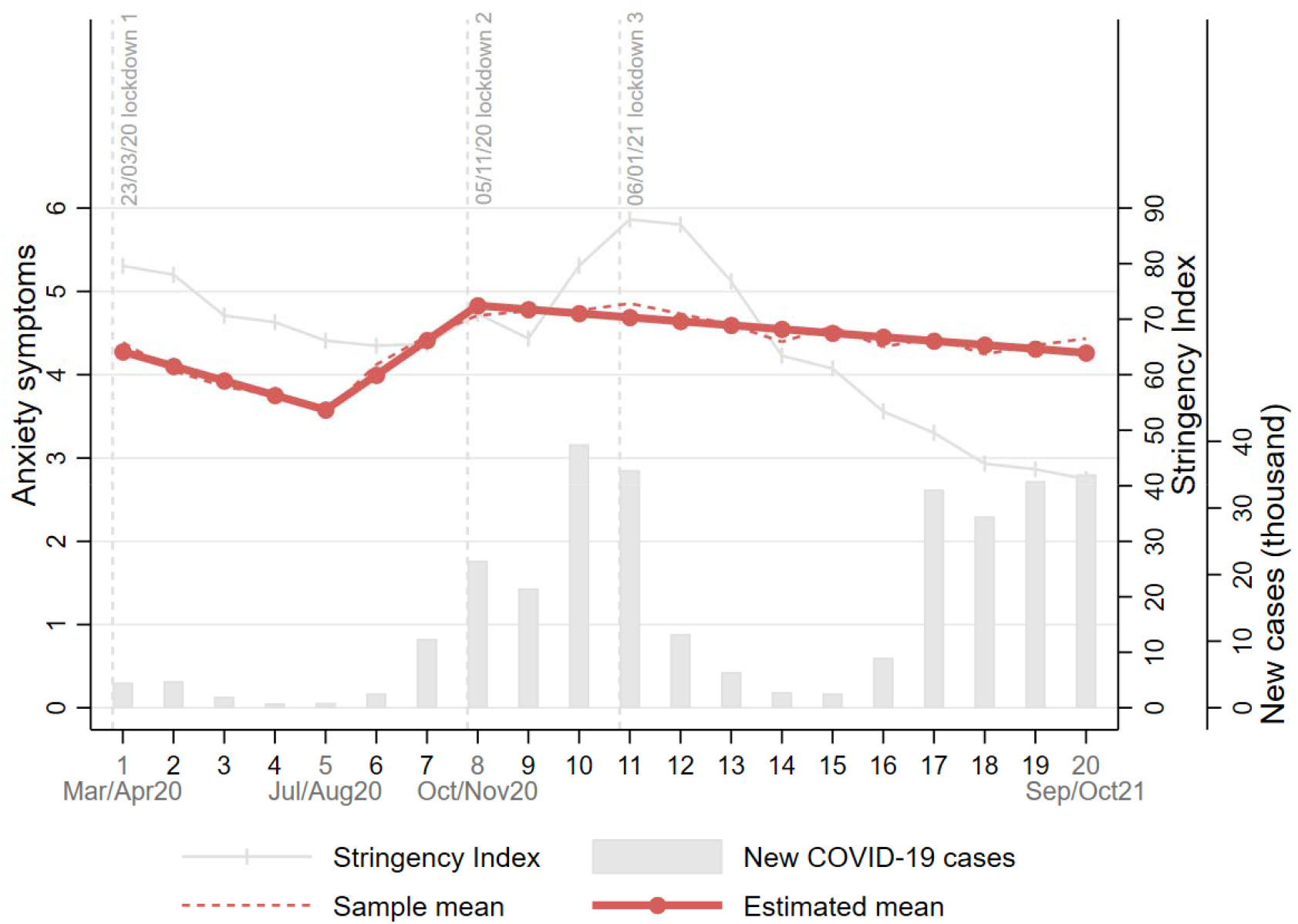
Overall growth trajectories of anxiety symptoms (unconditional model)

Results from the conditional LGMs are presented in Table 2. Compared with people living in an area of 10% greenspace or under, those with higher greenspace coverage had lower levels of anxiety at the start of the follow-up, in particular after controlling for all potential confounders (Model III). There was no evidence that greenspace coverage was related to the growth rate of anxiety during the period of first lockdown. However, there was some indication of an association with the rate of change at later stages of the pandemic. More specifically, people living in an area of 20 to 50% greenspace had a lower rate of increasing levels of anxiety between the first and second lockdown. This is depicted in Figure 2, which shows the estimated growth trajectories of anxiety symptoms by greenspace access categories based on the full model.

**Table 2.** Results from the latent growth models (N=19,848)

|  | Model I |  |  | Model II |  |  | Model III |  |  |
| --- | --- | --- | --- | --- | --- | --- | --- | --- | --- |
|  | Coef. | SE | p | Coef. | SE | p | Coef. | SE | p |
| **Greenspace:** |  |  |  |  |  |  |  |  |  |
| Intercept |  |  |  |  |  |  |  |  |  |
| <10-20% (vs. ≤10%) | **-0.50** | **0.19** | **0.007** | **-0.49** | **0.20** | **0.017** | **-0.54** | **0.18** | **0.003** |
| <20-50% (vs. ≤10%) | -0.11 | 0.19 | 0.539 | -0.14 | 0.23 | 0.539 | **-0.42** | **0.19** | **0.026** |
| >50% (vs. ≤10%) | **-0.55** | **0.18** | **0.002** | -0.52 | 0.29 | 0.078 | **-0.72** | **0.24** | **0.003** |
| Slope 1 |  |  |  |  |  |  |  |  |  |
| <10-20% (vs. ≤10%) | 0.03 | 0.03 | 0.331 | 0.04 | 0.04 | 0.243 | 0.04 | 0.04 | 0.239 |
| <20-50% (vs. ≤10%) | 0.02 | 0.04 | 0.590 | 0.04 | 0.04 | 0.410 | 0.04 | 0.04 | 0.304 |
| >50% (vs. ≤10%) | 0.01 | 0.03 | 0.848 | 0.03 | 0.05 | 0.566 | 0.04 | 0.05 | 0.491 |
| Slope 2 |  |  |  |  |  |  |  |  |  |
| <10-20% (vs. ≤10%) | -0.03 | 0.04 | 0.391 | -0.05 | 0.04 | 0.190 | -0.05 | 0.04 | 0.167 |
| <20-50% (vs. ≤10%) | -0.04 | 0.03 | 0.175 | -0.08 | 0.04 | 0.051 | **-0.09** | **0.04** | **0.024** |
| >50% (vs. ≤10%) | -0.03 | 0.04 | 0.446 | -0.06 | 0.05 | 0.247 | -0.07 | 0.05 | 0.199 |
| Slope 3 |  |  |  |  |  |  |  |  |  |
| <10-20% (vs. ≤10%) | 0.01 | 0.01 | 0.683 | 0.01 | 0.01 | 0.328 | 0.01 | 0.01 | 0.287 |
| <20-50% (vs. ≤10%) | 0.00 | 0.01 | 0.698 | 0.01 | 0.01 | 0.442 | 0.01 | 0.01 | 0.558 |
| >50% (vs. ≤10%) | 0.01 | 0.01 | 0.142 | 0.02 | 0.02 | 0.191 | 0.02 | 0.02 | 0.138 |
| **Growth factor:** |  |  |  |  |  |  |  |  |  |
| Intercept | **4.46** | **0.10** | **0.000** | **5.85** | **0.33** | **0.000** | **5.97** | **0.41** | **0.000** |
| Slope 1 | **-0.18** | **0.02** | **0.000** | **-0.18** | **0.06** | **0.001** | **-0.21** | **0.09** | **0.025** |
| Slope 2 | **0.44** | **0.02** | **0.000** | **0.57** | **0.06** | **0.000** | **0.59** | **0.10** | **0.000** |
| Slope 3 | **-0.05** | **0.01** | **0.000** | **-0.07** | **0.02** | **0.000** | **-0.07** | **0.03** | **0.018** |
| **Model fit:** |  |  |  |  |  |  |  |  |  |
| RMSEA<sup>†</sup> | 0.01 |  |  | 0.01 |  |  | 0.01 |  |  |
| CFI<sup>†</sup> | 0.99 |  |  | 0.99 |  |  | 0.99 |  |  |
| TLI<sup>†</sup> | 0.98 |  |  | 0.98 |  |  | 0.98 |  |  |
| SRMR<sup>†</sup> | 0.01 |  |  | 0.01 |  |  | 0.01 |  |  |

Notes: Model I, no covariate; Model II, controlling for population density and index of multiple deprivation; Model III, additionally controlling for individual characteristics; <sup>†</sup>RMSEA, Root Mean Square Error of Approximation; CFI, Comparative Fit Index; TLI, Tucker-Lewis Index; SRMR, Standardised Root Mean Square Residual

**Figure 2.**
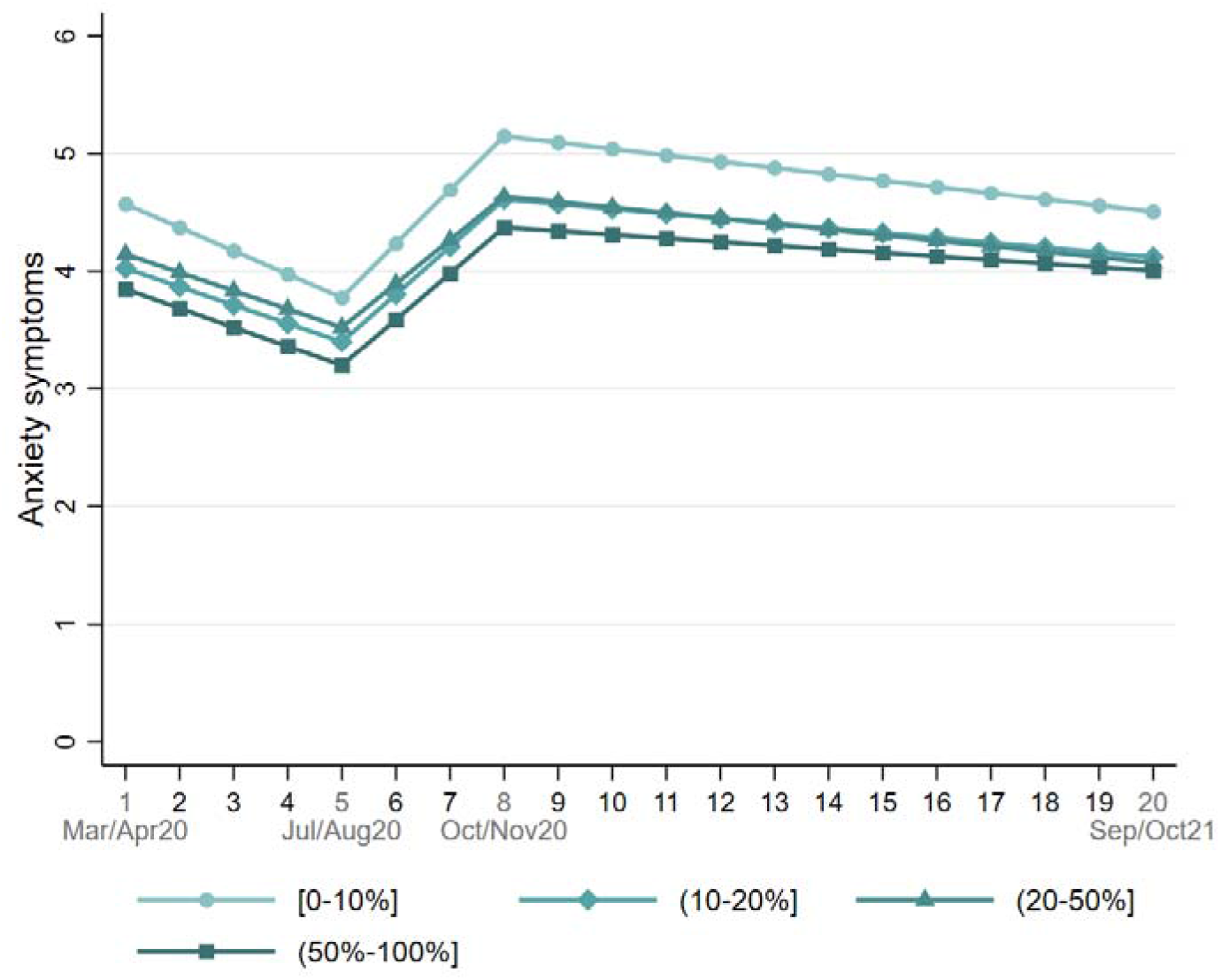
Estimated growth trajectories of anxiety symptoms by greenspace access (Model III)

### Sensitivity analyses

To test robustness of the results after taking into account greenspace satisfaction, we fitted LGMs based on a subsample with information on satisfaction with the availability of greenspace in the neighbourhood. The results are presented in Table S1 in the Supplementary Material. Being satisfied with greenspace was associated with fewer anxiety symptoms at baseline, but there was no evidence it was associated with the rate of change over time. Even after controlling for greenspace satisfaction, the objective greenspace measure was still found to be associated with both the intercept and rate of change of anxiety symptoms.

The more flexible free time scores approach had poorer model fit compared to the piecewise LGM (see Figure S2 in the Supplementary Material). There was no evidence that greenspace access measured by proximity was related to the growth trajectory of anxiety symptoms (Table S2). Finally, even after controlling for going outside of the house, the relationship between greenspace access and anxiety symptoms persisted (see Table S3).

## Discussion

This study examined how access to urban greenspace was related the growth trajectory of anxiety symptoms during the COVID-19 pandemic. Our results showed that living in an area with higher greenspace coverage was independently associated with fewer anxiety symptoms consistently across the 20-month observational period between March 2020 and October 2021. There was some evidence for the association of greenspace with the change of anxiety symptoms across different stages of the pandemic, but the differences in rate of change were relatively small.

Our findings are consistent with pre-pandemic studies^1,2,4^ and some of the recent studies during the COVID-19 pandemic^20,21^ showing potential benefits of greenspace for anxiety. It is promising that greenspace exposure was still shown to be associated with lower anxiety symptoms during the COVID-19 pandemic when there was a heightened risk of anxiety and other mental health conditions. Benefits may arise through a number of mechanisms, as described in the introduction. According to the stress reduction theory, exposure to greenspace could improve mental health through expediting recovery from stress.^11^ This is further supported by a recent meta-analysis showing that greenspace exposure is associated with decreased salivary cortisol levels which is a biomarker of psychological stress.^39^ This could be particularly relevant and salient during the pandemic due to increased stresses overall (e.g. bereavement, relationship breakdown, job loss) and reduced access to other mental health resource and support.^40^ In addition to stress, greenspace exposure might alleviate anxiety symptoms via other health promoting activities or behaviours. For example, greenspace (e.g. parks) could serve as social gathering locations when indoor activities were prohibited or discouraged during the pandemic.^41^ This was supported by evidence showing that people used greenspace as a way to maintain social interactions,^42,43^ which have been proven to be beneficial for mental health.^44^ In this regard, people living in an area with higher greenspace coverage are in an advantaged position. Moreover, the availability of greenspace was found to encourage outdoor activity more generally, in particular physical activity.^45^ Not only does it provide mental health benefits by changing the scenery and/or being away from stressors, but also exercise is suggested to have anxiolytic effects via physiological mechanisms, such as sympathetic nervous system and hypothalamic-pituitary-adrenal axis reactivity.^46^ The association between greenspace and outdoor activity is supported by a recent US study showing that human mobility reduction was lower in communities with better greenspace access during the early stage of the COVID-19 pandemic.^47^

Our sensitivity analyses additionally showed that the relationship between greenspace and anxiety symptoms remained even after accounting for going outdoors. In other words, the mental health benefits of greenspace were not explained away by the possibility that people living in areas with higher greenspace coverage might go out more. This is important to note, as the benefits of nature come not only from intentional interactions (e.g. visiting a park for recreation), but also from incidental (e.g. walking to work by a park) or indirect interactions (e.g. nature views at home).^48^ As a result of these direct and indirect ways of interacting with greenspace, patterns of different types of exposure were likely to vary across different stages of the COVID-19 pandemic related to the changes in virus containment policies. The predicted anxiety trajectories suggested that the difference in anxiety symptoms between people living in the lowest and highest greenspace coverage areas tended to be larger when COVID-19 policies were more stringent, and smaller when restrictions were relaxed. This is line with the fact that people were generally more restricted to their local communities during lockdowns, whereas the gap in greenspace exposure due to residence was mitigated due to human mobility (either intentional or incidental interactions) once lockdown measures were eased.

We recognise that it is challenging to draw any causal inference in observational studies. One challenge is to properly control for confounders. When it comes to the relationship between greenspace and mental health, one of the most important factors is arguably socioeconomic position (SEP). People from disadvantaged backgrounds are typically more restricted in their access to greenspace (private or public), due to either quantity or quality.^49^ At the same time, they tend to be at a higher risk of mental health problems.^50^ However, our analyses had controlled for relevant covariates at both individual and area levels, including income, education, population density and area deprivation. Thus, the observed association between greenspace and anxiety in this study could not be simply attributed to confounding by SEP.

This study has a number of strengths. It utilised a large sample with sufficient heterogeneity to include good stratification across all major socio-demographic groups and good coverage of geographic areas in England. The analyses were weighted on the basis of population estimates of core demographics, with the weighted data showing good alignment with a nationally representative study.^52^ The availability of postcode information allowed us to obtain greenspace coverage at small geographic areas (LSOA), controlling for other relevant geographic factors, including population density and area deprivation. Due to the longitudinal design of the COVID-19 Social Study, we were able to examine the growth trajectories of anxiety symptoms since the first lockdown in the UK across different stages of the COVID-19 pandemic over 20 months. Despite these strengths, the limitations of our study raise important points for further research. First, our data were from a non-probability sample. Despite the effort to make our sample representative to the population in England by weighting, there is still the possibility of potential biases due to omitting other demographic factors that could be associated with survey participation in the weighting process. Second, there is a lack of pre-pandemic data. It therefore remains unclear how the mental health benefits of greenspace in the context of COVID-19 pandemic compared to a normal scenario. Future research is encouraged to examine the mental health benefits of greenspace using data collected both prior to and during the pandemic.

The COVID-19 pandemic has had a profound effect on mental health and led to a sharp increase in demand for mental health assistance and interventions, presenting an unprecedented challenge to the National Health Service (NHS) in England. Greenspace is increasingly being recognised as an important asset for supporting mental health by policy makers and practitioners. In 2020, England launched a £5.77 million project on green social prescribing to prevent and tackle mental ill health. The recent Levelling Up White Paper included making greenspace accessible to all as one of its missions, by enhancing and maintaining green belts, parks, woodlands, particularly in communities with the lowest greenspace access.^53^ Our study showed that anxiety levels were consistently lower throughout the pandemic in areas with higher levels of greenspace, independent of other individual and geographical factors. This highlights the value of long-term investments in urban green infrastructure planning, as well as in improvement and maintenance of existing greenspace as a way of improving public mental health. Equally important is to raise public awareness of the mental health benefits of greenspace, and to facilitate and support engagement with greenspace especially among disadvantaged groups as an opportunity to tackle mental health inequality.

## Data Availability

Anonymous data will be made publicly available following the end of the pandemic.

## Declarations

### Ethics approval and consent to participate

The study was approved by the UCL Research Ethics Committee [12467/005] and all participants gave informed consent.

### Availability of data and materials

Anonymous data will be made publicly available following the end of the pandemic.

### Author contributions

FB, AS and DF developed the study idea and the analysis plan. BW derived the land cover metric. FB analysed the data. FB and HWM wrote the first draft. FB, HWM and DF had accessed and verified the underlying data. All authors provided critical revisions, read and approved the submitted manuscript.

## Acknowledgements

The researchers are grateful for the support of a number of organisations with their recruitment efforts including: the UKRI Mental Health Networks, Find Out Now, UCL BioResource, SEO Works, FieldworkHub, and Optimal Workshop. The study was also supported by HealthWise Wales, the Health and Care Research Wales initiative, which is led by Cardiff University in collaboration with SAIL, Swansea University.

## Reference

1. Collins, R. M. et al. A systematic map of research exploring the effect of greenspace on mental health. Landscape and Urban Planning 201, 103823 (2020).

2. Wendelboe-Nelson, C., Kelly, S., Kennedy, M. & Cherrie, J. W. A Scoping Review Mapping Research on Green Space and Associated Mental Health Benefits. International Journal of Environmental Research and Public Health 2019, Vol. 16, Page 2081 16, 2081 (2019).

3. Kotera, Y., Richardson, M. & Sheffield, D. Effects of Shinrin-Yoku (Forest Bathing) and Nature Therapy on Mental Health: a Systematic Review and Meta-analysis. International Journal of Mental Health and Addiction 1–25 (2020) doi:10.1007/S11469-020-00363-4/TABLES/6.

4. Gascon, M. et al. Mental Health Benefits of Long-Term Exposure to Residential Green and Blue Spaces: A Systematic Review. International Journal of Environmental Research and Public Health 2015, Vol. 12, Pages 4354-4379 12, 4354–4379 (2015).

5. Maas, J. et al. Morbidity is related to a green living environment. Journal of Epidemiology & Community Health 63, 967–973 (2009).

6. Astell-Burt, T. & Feng, X. Association of Urban Green Space With Mental Health and General Health Among Adults in Australia. JAMA Network Open 2, e198209–e198209 (2019).

7. Krekel, C., Kolbe, J. & Wüstemann, H. The greener, the happier? The effect of urban land use on residential well-being. Ecological Economics 121, 117–127 (2016).

8. Morita, E. et al. Psychological effects of forest environments on healthy adults: Shinrin-yoku (forest-air bathing, walking) as a possible method of stress reduction. Public Health 121, 54–63 (2007).

9. South, E. C., Hohl, B. C., Kondo, M. C., MacDonald, J. M. & Branas, C. C. Effect of Greening Vacant Land on Mental Health of Community-Dwelling Adults: A Cluster Randomized Trial. JAMA Network Open 1, e180298–e180298 (2018).

10. Kaplan, S. The restorative benefits of nature: Toward an integrative framework. Journal of Environmental Psychology 15, 169–182 (1995).

11. Ulrich, R. S. et al. Stress recovery during exposure to natural and urban environments. Journal of Environmental Psychology 11, 201–230 (1991).

12. Duncan, D. T. & Kawachi, I. Neighborhoods and Health. (Oxford University Press, 2018).

13. Markevych, I. et al. Exploring pathways linking greenspace to health: Theoretical and methodological guidance. Environmental Research 158, 301–317 (2017).

14. Hadjidemetriou, G. M., Sasidharan, M., Kouyialis, G. & Parlikad, A. K. The impact of government measures and human mobility trend on COVID-19 related deaths in the UK. Transportation Research Interdisciplinary Perspectives 6, 100167 (2020).

15. Geng, D. (Christina), Innes, J., Wu, W. & Wang, G. Impacts of COVID-19 pandemic on urban park visitation: a global analysis. Journal of Forestry Research 32, 553–567 (2021).

16. Venter, Z. S., Barton, D. N., Gundersen, V., Figari, H. & Nowell, M. Urban nature in a time of crisis: recreational use of green space increases during the COVID-19 outbreak in Oslo, Norway. Environmental Research Letters 15, 104075 (2020).

17. Pierce, M. et al. Mental health before and during the COVID-19 pandemic: a longitudinal probability sample survey of the UK population. The Lancet Psychiatry 7, 883–892 (2020).

18. Shanahan, L. et al. Emotional distress in young adults during the COVID-19 pandemic: evidence of risk and resilience from a longitudinal cohort study. Psychological Medicine 1–10 (2020) doi:10.1017/S003329172000241X.

19. Labib, S. M., Browning, M. H. E. M., Rigolon, A., Helbich, M. & James, P. Nature’s contributions in coping with a pandemic in the 21st century: A narrative review of evidence during COVID-19. Science of The Total Environment 833, 155095 (2022).

20. Pouso, S. et al. Contact with blue-green spaces during the COVID-19 pandemic lockdown beneficial for mental health. Science of The Total Environment 756, 143984 (2021).

21. Soga, M. et al. A room with a green view: the importance of nearby nature for mental health during the COVID-19 pandemic. Ecological Applications 31, e2248 (2021).

22. Heo, S., Desai, M. U., Lowe, S. R. & Bell, M. L. Impact of changed use of greenspace during covid-19 pandemic on depression and anxiety. International Journal of Environmental Research and Public Health 18, 5842 (2021).

23. Wortzel, J. D. et al. Association Between Urban Greenspace and Mental Wellbeing During the COVID-19 Pandemic in a U.S. Cohort. Frontiers in Sustainable Cities 3, 56 (2021).

24. Schindler, M., le Texier, M. & Caruso, G. How far do people travel to use urban green space? A comparison of three European cities. Applied Geography 141, 102673 (2022).

25. Ugolini, F., Massetti, L., Pearlmutter, D. & Sanesi, G. Usage of urban green space and related feelings of deprivation during the COVID-19 lockdown: Lessons learned from an Italian case study. Land Use Policy 105, 105437 (2021).

26. Natural England. The People and Nature Survey for England: Data and publications from Adults survey year 1 (April 2020 - March 2021). https://www.gov.uk/government/statistics/the-people-and-nature-survey-for-england-data-and-publications-from-adults-survey-year-1-april-2020-march-2021-official-statistics/the-people-and-nature-survey-for-england-data-and-publications-from-adults-survey-year-1-april-2020-march-2021-official-statistics-main-finding (2021).

27. Anxiety UK. Key Facts and Figures. https://www.anxietyuk.org.uk/media-centre/ (2020).

28. O’Shea, N. Forecasting needs and risks in the UK. Centre for Mental health https://www.centreformentalhealth.org.uk/publications/covid-19-and-nations-mental-health-october-2020 (2020).

29. HM Government. A Green Future: Our 25 Year Plan to Improve the Environment. https://www.gov.uk/government/publications/25-year-environment-plan (2018).

30. Alcock, I. et al. What accounts for ‘England’s green and pleasant land’? A panel data analysis of mental health and land cover types in rural England. Landscape and Urban Planning 142, 38–46 (2015).

31. Spitzer, R. L., Kroenke, K., Williams, J. B. W. & Löwe, B. A Brief Measure for Assessing Generalized Anxiety Disorder. Archives of Internal Medicine 166, 1092 (2006).

32. Morton, R. D., Marston, C. G., O’Neil, A. W. & Rowland, C. S. Land Cover Map 2019 (20m classified pixels, GB). https://doi.org/10.5285/643eb5a9-9707-4fbb-ae76-e8e53271d1a0 (2020).

33. Wheeler, B. W. et al. Beyond greenspace: An ecological study of population general health and indicators of natural environment type and quality. International Journal of Health Geographics 14, 1–17 (2015).

34. Office for National Statistics. Access to gardens and public green space in Great Britain. https://www.ons.gov.uk/economy/environmentalaccounts/datasets/accesstogardensandpublicgreenspaceingreatbritain.

35. Fancourt, D., Bu, F., Mak, H. W., Paul, E. & Steptoe, A. Covid-19 Social Study: Results Release 40. https://www.covidsocialstudy.org/_files/ugd/064c8b_86930bad37754dc9ac0553ef44caa902.pdf (2021).

36. Fancourt, D., Steptoe, A. & Bu, F. Trajectories of anxiety and depressive symptoms during enforced isolation due to COVID-19 in England: a longitudinal observational study. The Lancet Psychiatry 8, 141–149 (2021).

37. Office for National Statistics. Population estimates for the UK, England and Wales, Scotland and Northern Ireland. https://www.ons.gov.uk/peoplepopulationandcommunity/populationandmigration/populationestimates/bulletins/annualmidyearpopulationestimates/mid2018 (2019).

38. Hale, T. et al. A global panel database of pandemic policies (Oxford COVID-19 Government Response Tracker). Nature Human Behaviour 2021 5:4 5, 529–538 (2021).

39. Twohig-Bennett, C. & Jones, A. The health benefits of the great outdoors: A systematic review and meta-analysis of greenspace exposure and health outcomes. Environmental Research 166, 628–637 (2018).

40. WHO. Action required to address the impacts of the COVID-19 pandemic on mental health and service delivery systems in the WHO European Region: recommendations from the European Technical Advisory Group on the mental health impacts of COVID-19, 30 June 2021. https://www.euro.who.int/en/health-topics/noncommunicable-diseases/mental-health/publications/2021/action-required-to-address-the-impacts-of-the-covid-19-pandemic-on-mental-health-and-service-delivery-systems-in-the-who-european-region-recommendations-from-the-european-technical-advisory-group-on-the-mental-health-impacts-of-covid-19,-30-june-2021 (2021).

41. Mouratidis, K. How COVID-19 reshaped quality of life in cities: A synthesis and implications for urban planning. Land Use Policy 111, 105772 (2021).

42. McCormack, G. R., Petersen, J., Naish, C., Ghoneim, D. & Doyle-Baker, P. K. Neighbourhood environment facilitators and barriers to outdoor activity during the first wave of the COVID-19 pandemic in Canada: a qualitative study. https://doi.org/10.1080/23748834.2021.2016218 (2022) doi:10.1080/23748834.2021.2016218.

43. Borkenhagen, D. et al. The effect of COVID-19 on parks and greenspace use during the first three months of the pandemic – a survey study. https://doi.org/10.1080/23748834.2021.1963646 (2021) doi:10.1080/23748834.2021.1963646.

44. Kawachi, I. & Berkman, L. F. Social ties and mental health. Journal of Urban Health 2001 78:3 78, 458–467 (2001).

45. Kondo, M. C., Fluehr, J. M., McKeon, T. & Branas, C. C. Urban Green Space and Its Impact on Human Health. International Journal of Environmental Research and Public Health 2018, Vol. 15, Page 445 15, 445 (2018).

46. Anderson, E. & Shivakumar, G. Effects of exercise and physical activity on anxiety. Frontiers in Psychiatry 4, 27 (2013).

47. Heo, S., Lim, C. C. & Bell, M. L. Relationships between Local Green Space and Human Mobility Patterns during COVID-19 for Maryland and California, USA. Sustainability 2020, Vol. 12, Page 9401 12, 9401 (2020).

48. Keniger, L. E., Gaston, K. J., Irvine, K. N. & Fuller, R. A. What are the Benefits of Interacting with Nature? International Journal of Environmental Research and Public Health 2013, Vol. 10, Pages 913-935 10, 913–935 (2013).

49. Geary, R. S. et al. A call to action: Improving urban green spaces to reduce health inequalities exacerbated by COVID-19. Preventive Medicine 145, 106425 (2021).

50. Marmot, M., Allen, J., Boyce, T., Goldblatt, P. & Morrison, J. Health Equity in England: The Marmot Review 10 Years On - The Health Foundation. https://www.health.org.uk/publications/reports/the-marmot-review-10-years-on (2020).

51. Mitchell, R. & Popham, F. Effect of exposure to natural environment on health inequalities: an observational population study. The Lancet 372, 1655–1660 (2008).

52. Bu, F., Steptoe, A. & Fancourt, D. Who is lonely in lockdown? Cross-cohort analyses of predictors of loneliness before and during the COVID-19 pandemic. Public Health 186, 31–34 (2020).

53. HM Government. Levelling Up the United Kingdom. www.gov.uk/official-documents (2022).

