## Supplement for "Urban greenspace and anxiety symptoms during the COVID-19 pandemic: A 20-month follow up of 19,848 participants in England"

### Supplementary Material

Table S1 Results from the latent growth models with both quantity and quality measures of greenspace (N=12,570)

| | Model I | | | Model II | | | Model III | | |
|---|---|---|---|---|---|---|---|---|---|
| | Coef. | SE | p | Coef. | SE | p | Coef. | SE | p |
| **Greenspace:** | | | | | | | | | |
| Intercept | | | | | | | | | |
| <10-20% (vs. ≤10%) | **-0.70** | **0.21** | **0.001** | **-0.66** | **0.24** | **0.006** | **-0.60** | **0.22** | **0.006** |
| <20-50% (vs. ≤10%) | -0.09 | 0.24 | 0.698 | -0.11 | 0.29 | 0.695 | -0.29 | 0.25 | 0.238 |
| >50% (vs. ≤10%) | -0.35 | 0.21 | 0.097 | -0.45 | 0.35 | 0.198 | **-0.71** | **0.30** | **0.019** |
| Satisfied (vs. other) | **-1.38** | **0.27** | **<0.001** | **-1.21** | **0.26** | **<0.001** | **-0.91** | **0.22** | **<0.001** |
| Slope 1 | | | | | | | | | |
| <10-20% (vs. ≤10%) | 0.03 | 0.04 | 0.405 | 0.04 | 0.04 | 0.321 | 0.04 | 0.04 | 0.301 |
| <20-50% (vs. ≤10%) | 0.01 | 0.04 | 0.823 | 0.03 | 0.05 | 0.608 | 0.04 | 0.05 | 0.454 |
| >50% (vs. ≤10%) | 0.02 | 0.04 | 0.575 | 0.04 | 0.07 | 0.523 | 0.05 | 0.06 | 0.427 |
| Satisfied (vs. other) | -0.03 | 0.04 | 0.496 | -0.03 | 0.04 | 0.512 | -0.02 | 0.05 | 0.668 |
| Slope 2 | | | | | | | | | |
| <10-20% (vs. ≤10%) | 0.00 | 0.04 | 0.959 | -0.01 | 0.04 | 0.760 | -0.02 | 0.04 | 0.709 |
| <20-50% (vs. ≤10%) | -0.06 | 0.04 | 0.081 | -0.08 | 0.04 | 0.058 | **-0.09** | **0.04** | **0.032** |
| >50% (vs. ≤10%) | -0.01 | 0.05 | 0.894 | -0.03 | 0.06 | 0.652 | -0.04 | 0.06 | 0.480 |
| Satisfied (vs. other) | -0.05 | 0.04 | 0.212 | -0.04 | 0.04 | 0.297 | -0.03 | 0.04 | 0.457 |
| Slope 3 | | | | | | | | | |
| <10-20% (vs. ≤10%) | 0.01 | 0.01 | 0.576 | 0.02 | 0.01 | 0.121 | 0.02 | 0.01 | 0.104 |
| <20-50% (vs. ≤10%) | 0.00 | 0.01 | 0.920 | 0.02 | 0.01 | 0.104 | 0.02 | 0.01 | 0.110 |
| >50% (vs. ≤10%) | 0.01 | 0.01 | 0.217 | **0.04** | **0.02** | **0.018** | **0.04** | **0.02** | **0.016** |
| Satisfied (vs. other) | 0.00 | 0.01 | 0.954 | 0.00 | 0.01 | 0.972 | 0.00 | 0.01 | 0.881 |
| **Growth factor:** | | | | | | | | | |
| Intercept | **5.20** | **0.27** | **<0.001** | **6.13** | **0.44** | **<0.001** | **6.07** | **0.52** | **<0.001** |
| Slope 1 | **-0.20** | **0.04** | **<0.001** | **-0.22** | **0.07** | **0.002** | **-0.33** | **0.11** | **0.003** |
| Slope 2 | **0.46** | **0.04** | **<0.001** | **0.53** | **0.07** | **<0.001** | **0.52** | **0.11** | **<0.001** |
| Slope 3 | **-0.04** | **0.01** | **0.008** | **-0.07** | **0.02** | **<0.001** | -0.05 | 0.03 | 0.098 |
| **Model fit:** | | | | | | | | | |
| RMSEA† | 0.02 | | | 0.02 | | | 0.01 | | |
| CFI† | 0.98 | | | 0.98 | | | 0.98 | | |
| TLI† | 0.98 | | | 0.98 | | | 0.98 | | |
| SRMR† | 0.01 | | | 0.01 | | | 0.01 | | |

Notes: Model I, no covariate; Model II, controlling for population density and index of multiple deprivation; Model III, additionally controlling for individual characteristics; †RMSEA, Root Mean Square Error of Approximation; CFI, Comparative Fit Index; TLI, Tucker-Lewis Index; SRMR, Standardised Root Mean Square Residual

Table S2 Results from the latent growth models with alternative measure of greenspace (N=19,848)

| | Model III-I | | | Model III-II | | |
|---|---|---|---|---|---|---|
| | Coef. | SE | p | Coef. | SE | p |
| **Greenspace:** | | | | | | |
| Intercept | | | | | | |
| Proximity | 0.05 | 0.03 | 0.077 | 0.07 | 0.08 | 0.379 |
| Proximity$^2$ | | | | 0.00 | 0.01 | 0.801 |
| Slope 1 | | | | | | |
| Proximity | 0.00 | 0.01 | 0.543 | 0.00 | 0.02 | 0.847 |
| Proximity$^2$ | | | | 0.00 | 0.00 | 0.953 |
| Slope 2 | | | | | | |
| Proximity | **0.01** | **0.01** | **0.048** | 0.02 | 0.01 | 0.213 |
| Proximity$^2$ | | | | -0.00 | 0.00 | 0.627 |
| Slope 3 | | | | | | |
| Proximity | -0.00 | 0.00 | 0.713 | -0.01 | 0.01 | 0.338 |
| Proximity$^2$ | | | | 0.00 | 0.00 | 0.343 |
| **Growth factor:** | | | | | | |
| Intercept | **5.19** | **0.36** | **<0.001** | **5.15** | **0.38** | **<0.001** |
| Slope 1 | -0.15 | 0.09 | 0.089 | -0.16 | 0.10 | 0.100 |
| Slope 2 | **0.47** | **0.09** | **<0.001** | **0.46** | **0.10** | **<0.001** |
| Slope 3 | -0.05 | 0.03 | 0.082 | -0.04 | 0.03 | 0.161 |
| **Model fit:** | | | | | | |
| RMSEA$^†$ | 0.01 | | | 0.01 | | |
| CFI$^†$ | 0.99 | | | 0.99 | | |
| TLI$^†$ | 0.98 | | | 0.98 | | |
| SRMR$^†$ | 0.01 | | | 0.01 | | |

Notes: $^†$RMSEA, Root Mean Square Error of Approximation; CFI, Comparative Fit Index; TLI, Tucker-Lewis Index; SRMR, Standardised Root Mean Square Residual

Table S3 Results from the latent growth models controlling for days of going outside (N=19,848)

| | Model IV | | |
|---|---|---|---|
| | Coef. | SE | p |
| Intercept | | | |
| <10-20% (vs. ≤10%) | **-0.54** | **0.18** | **0.003** |
| <20-50% (vs. ≤10%) | **-0.42** | **0.19** | **0.026** |
| >50% (vs. ≤10%) | **-0.72** | **0.24** | **0.003** |
| Slope 1 | | | |
| <10-20% (vs. ≤10%) | 0.04 | 0.04 | 0.269 |
| <20-50% (vs. ≤10%) | 0.04 | 0.04 | 0.322 |
| >50% (vs. ≤10%) | 0.03 | 0.05 | 0.558 |
| Slope 2 | | | |
| <10-20% (vs. ≤10%) | -0.06 | 0.04 | 0.157 |
| <20-50% (vs. ≤10%) | **-0.09** | **0.04** | **0.021** |
| >50% (vs. ≤10%) | -0.07 | 0.05 | 0.190 |
| Satisfied (vs. other) | | | |
| Slope 3 | | | |
| <10-20% (vs. ≤10%) | 0.01 | 0.01 | 0.262 |
| <20-50% (vs. ≤10%) | 0.01 | 0.01 | 0.552 |
| >50% (vs. ≤10%) | 0.03 | 0.02 | 0.117 |
| Going out 1 | 0.00 | 0.01 | 0.788 |
| Going out 2 | **-0.05** | **0.01** | **<0.001** |
| Going out 3 | **-0.06** | **0.01** | **<0.001** |
| Going out 4 | **-0.07** | **0.01** | **<0.001** |
| Going out 5 | **-0.09** | **0.01** | **<0.001** |
| Going out 6 | **-0.07** | **0.01** | **<0.001** |
| Going out 7 | **-0.08** | **0.01** | **<0.001** |
| Going out 8 | **-0.11** | **0.01** | **<0.001** |
| Going out 9 | **-0.09** | **0.01** | **<0.001** |
| Going out 10 | **-0.08** | **0.01** | **<0.001** |
| Going out 11 | **-0.08** | **0.01** | **<0.001** |
| Going out 12 | **-0.09** | **0.01** | **<0.001** |
| Going out 13 | **-0.10** | **0.01** | **<0.001** |
| Going out 14 | **-0.11** | **0.01** | **<0.001** |
| Going out 15 | **-0.08** | **0.01** | **<0.001** |
| Going out 16 | **-0.11** | **0.01** | **<0.001** |
| Going out 17 | **-0.09** | **0.01** | **<0.001** |
| Going out 18 | **-0.11** | **0.01** | **<0.001** |
| Going out 19 | **-0.09** | **0.01** | **<0.001** |
| Going out 20 | **-0.07** | **0.01** | **<0.001** |
| **Growth factor:** | | | |
| Intercept | **6.02** | **0.41** | **<0.001** |
| Slope 1 | -0.13 | 0.09 | 0.178 |
| Slope 2 | **0.59** | **0.10** | **<0.001** |
| Slope 3 | **-0.06** | **0.03** | **0.029** |

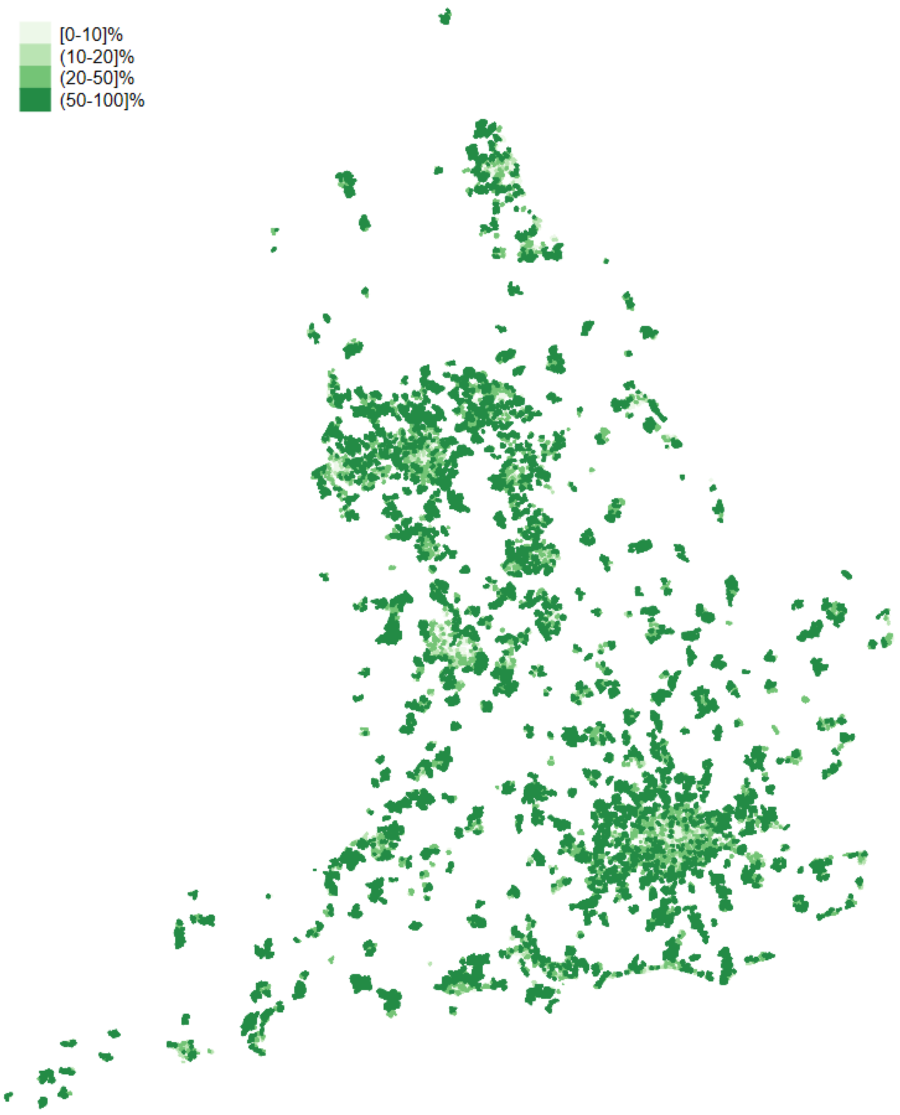


Notes: *Based upon Land Cover Map 2019 © UKCEH 2020. Contains Ordnance Survey data © Crown Copyright 2007, Licence number 100017572.*

Figure S1 Distribution of urban geographic areas that were included in the analyses (categorised by greenspace exposure)

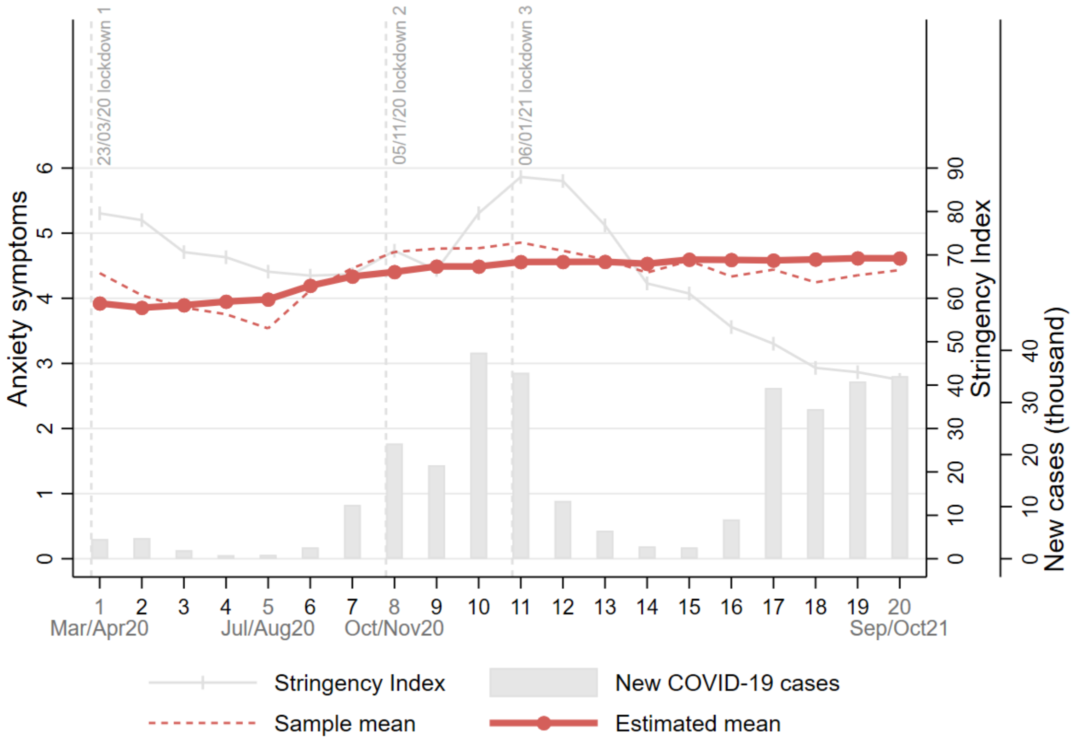


Notes: RMSEA=0.03, CFI=0.95, TLI=0.95, SRMR=0.03

Figure S2 Overall growth trajectories of anxiety symptoms (unconditional model with free time scores)
